# Memory perfectionism is associated with persistent memory complaints after concussion

**DOI:** 10.1101/2021.11.15.21266362

**Authors:** Edwina L. Picon, Evgenia Todorova, Daniela J. Palombo, David L. Perez, Andrew Howard, Noah D. Silverberg

**Author notes:** Corresponding author: Dr. Noah D. Silverberg, Department of Psychology, University of British Columbia, 3505- 2136 West Mall, Vancouver, British Columbia, Canada, V6T 1Z4. Disclosure of relationships and activities:* D.L.P. receives honoraria for continuing medical education lectures on functional neurological disorder. A.H. receives fees for medical-legal consulting involving patients with concussion. N.D.S. receives honoraria for continuing medical education lectures on concussion and fees for medical-legal and clinical neuropsychological consultation.

## Abstract

**Objective:** The etiology of persistent memory complaints after concussion is poorly understood. Memory perfectionism (highly valuing memory ability and intolerance of minor memory lapses) may help explain why some people report persistent subjective memory problems in the absence of corresponding objective memory impairment. The present study investigated the relationship between memory perfectionism and persistent memory complaints after concussion.

**Methods:** Adults (N=77; 61% women) with persistent symptoms following concussion were recruited from outpatient specialty clinics. Participants completed the National Institutes of Health Toolbox Cognition Battery, Test of Memory Malingering-Trial 1, and questionnaires measuring memory perfectionism (Memory in Adulthood-Achievement subscale), forgetfulness and other post-concussion symptoms (Rivermead Postconcussion Symptoms Questionnaire; RPQ), and depression (Patient Health Questionnaire-2) at M=17.8 weeks post-injury. Patients with vs. without severe memory complaints (based on the RPQ) were compared.

**Results:** Memory perfectionism was associated with severe memory complaint, after controlling for objective memory ability, overall cognitive ability, and depression (95% confidence interval for odds ratio = 1.11 to 1.40). Sensitivity analyses showed that this relationship did not depend on use of specific objective memory tests nor on inclusion of participants who failed performance validity testing. In a control comparison to test the specificity of identified relationships, memory perfectionism was not associated with severe fatigue (95% confidence interval for odds ratio = 0.91 to 1.07).

**Discussion:** Memory perfectionism may predispose people to experience persistent memory symptoms and/or contribute to their perpetuation after concussion, with potential relevance to the spectrum of functional cognitive disorders more broadly.

## INTRODUCTION

Most people who sustain a concussion demonstrate normal cognitive function on neuropsychological testing by three months after injury^1^, however, almost half continue to subjectively experience cognitive problems for a year or longer, with memory complaints being one of the most common^2–4^. The etiology of these complaints is not clear. The relationship between cognitive complaints and objective cognitive performance on neuropsychological testing, cross-sectionally or longitudinally, is weak or not significant in most studies^5–9^. The minority of studies that reported an association between subjective and objective cognitive functioning on testing did not control for symptom or performance validity, which could have affected their results^10,11^. Persistent cognitive complaints after concussion also correlate weakly with indicators of injury severity^8^, trauma-related macrostructural brain lesions^4,12^ and white matter integrity^13–15^. Stronger correlates of cognitive complaints after concussion include pre-injury history of psychiatric conditions^10^, personality traits such as predisposition to interoceptive focus^12^, and current symptoms of depression, anxiety and post-traumatic stress^5,7,16^. In summary, underlying cognitive impairment is probably not the primary basis for persistent cognitive symptoms after concussion.

In many individuals with persistent cognitive symptoms after concussion, the clinical presentation is consistent with Functional Cognitive Disorder (FCD), a clinical entity characterized by an internal inconsistency between subjective cognitive symptoms and objective cognitive performance^17,18^. The FCD literature may help generate hypotheses about why cognitive symptoms (most commonly relating to memory) often persist after concussion in the absence of underlying cognitive impairment. In a recently proposed theoretical model of FCD across various clinical settings^19^, memory perfectionism was positioned as a key predisposing and perpetuating factor. Memory perfectionism refers to the combination of a strongly valued memory ability and an intolerance for minor memory lapses^19^. Memory perfectionism is thought to promote self-monitoring, increase noticing of memory lapses, and incite catastrophic interpretations of benign lapses (e.g., as evidence of permanent brain damage), all of which reinforces attention to and concern for memory problems^19,20^.

Memory perfectionism correlates with the severity of memory complaints in people who have no objective impairment on memory testing, in both community and clinical samples^21,22^. To date, no studies have investigated memory perfectionism in the context of post-concussion symptoms. The objective of the present cohort study was to explore the association between memory perfectionism and persistent memory complaints following concussion. We hypothesized that higher memory perfectionism would be uniquely associated with memory complaint severity, after controlling for potential confounds, including objective episodic memory ability, global cognitive ability, and depression. Furthermore, through sensitivity analyses, we aimed to demonstrate that low effort in a subset of participants could not account for these relationships and that memory perfectionism is not similarly associated with severity of post-concussion symptoms other than memory (e.g., fatigue).

## METHODS

The present study is a secondary analysis of baseline data from a randomized control trial (Clinicaltrials.gov NCT03972579) as well as participants who were ineligible for this trial because they did not score highly on measures of maladaptive coping. Prior to randomization, participants completed an in-person assessment with a research assistant, under the supervision of a board-certified neuropsychologist (NDS). The assessment included self-report measures and neuropsychological testing. The University of British Columbia Behavioural Research Ethics Board and the Vancouver Coastal Health Research Institute Research Ethics Boards provided ethics approval for this study.

### Participants

Informed consent was obtained from all participants before starting the baseline assessment. Participants recruited after COVID-19 restrictions came into effect (March 14, 2020) were excluded from the present study because they completed a modified online baseline assessment that did not include neuropsychological testing. Seventy-seven participants were recruited from two multidisciplinary, public sector outpatient concussion clinics in British Columbia, Canada between April 2019 and March 2020. Patients were eligible for inclusion in the present study if they (1) were aged 18-69 years old, (2) sustained a concussion (mild traumatic brain injury) according to the World Health Organization Neurotrauma Task Force definition^23^ between 1 and 12 months prior to recruitment, (3) were fluent in English, and (4) self-reported ≥ 3 moderate-severe symptoms on the Rivermead Postconcussion Symptoms Questionnaire (RPQ) ^24^, a commonly used cut-off for “symptomatic” status^2^.

### Measures

#### Rivermead Post Concussion Symptoms Questionnaire (RPQ) ^24^

The RPQ is a widely used self-report measure of symptom severity in concussion research and is recommended in the National Institute of Neurological Disorders and Stroke Common Data Elements for traumatic brain injury and sport-related concussion. Participants rate a list of 16 symptoms on scale ranging from 0 (not experienced at all) to 4 (severe problem), in comparison to before their concussion. Subjective memory complaints were measured with the “forgetfulness” item on the RPQ.

#### Metamemory in Adulthood Questionnaire (MIA)^25^

As in a prior FCD study^21^, we measured memory perfectionism with the Achievement subscale from the MIA ^25^. The Achievement subscale consists of 16 items such as “It’s important that I am very accurate when remembering names of people” and “It bothers me when others notice my memory failures,” which are rated on a scale of 1 (agree strongly) to 5 (disagree strongly). After reverse scoring certain items, higher total MIA-Achievement scores reflect higher self-perceived memory perfectionism. In healthy people, intraindividual differences on the MIA-Achievement scale are stable through adulthood, supporting memory perfectionism as a personality trait^26,27^.

#### Patient Health Questionnaire-2 (PHQ-2)^28^

We assessed depression with the Patient Health Questionnaire (PHQ-9), a reliable and valid self-report measure of depression symptom severity over the preceding two weeks^29^. The PHQ-9 includes items that overlap with the RPQ (e.g., difficulty concentrating and fatigue). The minimize this overlap, we analyzed the PHQ-2 in this study, which includes only the two cardinal depression symptoms (sadness and anhedonia) and has demonstrated accuracy for detecting depression that rivals the full PHQ-9^28^.

#### National Institutes of Health Toolbox Cognition Battery (NIHTB-CB)^30^

The NIHTB-CB is administered by a trained examiner with the iPad and includes standardized measures of cognitive abilities. The Picture Sequence Memory Test, which requires participants to reproduce the sequence of pictures briefly presented is the core episodic memory test in the NIHTB-CB and for the present study. It has demonstrated good test-retest reliability and expected associations with “gold standard” memory tests and health-related variables (e.g., age)^31^. We used the NIHTB-CB Total Cognition Composite score as an index of global cognitive ability. It is comprised of the Picture Sequence Memory Test and the six other core tests from the NIHTB-CB assessing crystallized intelligence, executive function, processing speed, and attention/working memory. Participants in this study were also administered the abbreviated Rey Auditory Verbal Learning Test (RAVLT), created as a supplementary measure for, and co-normed with, the NIHTB-CB. In this task, participants hear a list of fifteen words and then must repeat as many of the words as they can. The NIHTB-CB RAVLT score is the sum of correctly recalled words over three consecutive learning trials. We considered the NIHTB-CB RAVLT as alternative measure of episodic memory in sensitivity analyses.

#### Test of Memory Malingering^32^

We measured performance validity using Trial 1 of the Test of Memory Malingering (TOMM-1). The TOMM-1 has the appearance of a challenging (visual recognition) memory test but was designed as a test of *intention* to perform well, i.e., effort or performance validity. In a recent meta-analysis, a cut-off score of below 42 on the TOMM-1 optimally identifies test-takers with known performance invalidity (sensitivity=0.6-0.7 and specificity>0.9)^33^.

### Statistical Analysis

Statistical analysis was performed using SPSS version 27 (SPSS, Chicago, IL, USA). Our primary outcome was subjective memory complaints as measured by the “forgetfulness” item on the RPQ. We dichotomized this ordinal scale into a binary variable reflecting the presence (item score=4) or absence (item score=0-3) of severe memory problems, because (1) a high proportion of participants (n=27 out of 77; 35%) endorsed severe memory problems, perhaps not surprising for patients seeking specialty treatment for persistent symptoms, (2) certain cell sizes were insufficient for analysis (only 3 of 77 participants endorsed an item score of 0), and (3) the mean item score for all items on the RPQ was 2.8; in the context of a high overall symptom burden, we wished to identify participants for whom memory was an area of particular clinical concern.

In all logistic regression models, the binary RPQ forgetfulness item was the dependent variable and MIA-Achievement (memory perfectionism) was the main independent variable. Additional covariates included episodic memory ability (NIHTB-CB Picture Sequence Memory Test), global cognitive ability (NIHTB-CB Total Cognition Composite) and depression (PHQ-2). We conducted several sensitivity analyses to assess the robustness of our primary findings. First, we substituted the NIHTB-CB RAVLT for the NIHTB-CB Picture Sequence Memory Test in logistic regression modeling to see if a different (auditory-verbal vs. visual) objective measure of episodic memory would alter our findings. Second, to determine if the relationship between memory perfectionism and symptom reporting was specific to memory complaints, we substituted the RPQ fatigue item (a non-memory symptom with a similar frequency distribution) for the forgetfulness item, converting it to binary in the same manner as was done for the forgetfulness item (4=presence of severe complaint, 0-3=absent). Finally, we ran a model excluding the 13 participants who scored below 42 on the TOMM-1 to determine if cases of performance invalidity accounted for the associations between memory perfectionism and memory complaints in the primary analysis.

## RESULTS

Participants’ baseline demographic and injury characteristics are presented in **Table 1**. The primary logistic regression model is presented in **Table 2**. Higher memory perfectionism (MIA-Achievement) was associated with severe memory complaint (odds ratio = 1.25, 95% confidence interval = 1.11 to 1.40, p < 0.001), when controlling for depression (PHQ-2), objective memory ability (NIHTB-CB Picture Sequence Memory), and global cognitive ability (NIHTB-CB Total Cognition Composite).

**Table 1.** Participant characteristics.

| **Variable** | **N** | **M (SD) / n (%)** |
| --- | --- | --- |
| Age, M (SD) | 77 | 42.35 (11.44) |
| Sex, n (% female) | 77 | 47 (61%) |
| Education level, n (%) | 77 |  |
| High school |  | 4 (5%) |
| Some college/university, no degree |  | 17 (22%) |
| Two-year college degree |  | 18 (23%) |
| Bachelor's degree |  | 20 (26%) |
| Graduate degree |  | 18 (24%) |
| Ethnicity, n (%) | 77 |  |
| White |  | 58 (75%) |
| Asian |  | 12 (16%) |
| Other |  | 7 (9%) |
| Weeks post injury, M (SD) | 77 | 17.74 (10.06) |
| Mechanism of injury, n (%) | 77 |  |
| Motor vehicle crash |  | 31 (40%) |
| Fall |  | 13 (17%) |
| Sport / recreational |  | 19 (25%) |
| Other |  | 14 (18%) |
| Loss of consciousness, n (%) | 77 |  |
| Yes |  | 10 (13%) |
| Suspected |  | 9 (12%) |
| No |  | 53 (69%) |
| Unknown |  | 5 (6%) |
| Co-occurring orthopedic injury, n (%) | 77 | 58 (75%) |
| Previous concussion, n (%) | 77 | 26 (34%) |
| Litigating, n (%) | 77 |  |
| Yes |  | 23 (30%) |
| No |  | 43 (56%) |
| Not applicable |  | 11 (14%) |
| PHQ-2, M (SD) | 77 | 2.61 (1.60) |
| MIA-Achievement, M (SD) | 77 | 67.34 (7.30) |
| RPQ total score, M (SD) | 77 | 35.74 (13.72) |
| NIHTB-CB Picture Sequence Memory, M (SD) | 76 | 99.74 (15.77) |
| NIHTB-CB RAVLT, M (SD) | 76 | 23.75 (6.04) |
| NIHTB-CB Total Cognition Composite, M (SD) | 76 | 100.75 (15.15) |

**Table 2.** Final logistic regression model.

|  | **B** | **Wald** | **Sig.** | **Odds ratio** | **95% CI Lower** | **95% CI Upper** |
| --- | --- | --- | --- | --- | --- | --- |
| MIA-Achievement | 0.222 | 12.659 | < 0.001 | 1.249 | 1.105 | 1.411 |
| NIHTB-CB Picture Sequence Memory Test | -0.009 | 0.126 | 0.722 | 0.991 | 0.945 | 1.040 |
| NIHTB-CB Total Cognition Composite | -0.012 | 0.312 | 0.577 | 0.988 | 0.946 | 1.032 |
| Patient Health Questionnaire-2 | 0.302 | 2.250 | 0.134 | 1.352 | 0.912 | 2.006 |

NIHTB-CB=National Institutes of Health Toolbox Cognition Battery; MIA=Metamemory in Adulthood Questionnaire.

Multiple sensitivity analyses were run to check the robustness of this finding.

Substituting the NIHTB-CB Picture Sequence Memory for the NIHTB-CB RAVLT resulted in a similar magnitude effect of memory perfectionism, OR = 1.242, 95% CI = 1.099-1.402, p < 0.001. When substituting the primary outcome (RPQ Forgetfulness) for a non-memory related symptom (RPQ Fatigue), memory perfectionism was no longer a significant predictor, OR = .985, 95% CI = .909 – 1.067, p = .712. Higher depression scores (PHQ-2) emerged as a significant predictor of fatigue in this model, OR = 1.880, 95% CI = 1.263-2.798, p = .002. Finally, to determine whether the association between memory perfectionism and severe memory complaint hold for only participants who passed performance validity testing, we removed the 13 participants who scored below 42 on the TOMM from analyses and re-ran the aforementioned regression models on the remaining 64 participants. Results were consistent with the previous models that included all participants. MIA Achievement was significantly associated with “severe” RPQ forgetfulness in the model with NIHTB-CB Picture Sequence Memory, OR = 1.303, 95% CI = 1.102-1.541, p = .002, and in the model with RAVLT, OR = 1.290, 95% CI = 1.091-1.526, p = .003. MIA-Achievement was not associated with RPQ Fatigue, OR = .974, 95% CI = .893-1.063, p = .560, however depression (PHQ-2) remained a significant predictor of fatigue, OR = 1.976, 95% CI = 1.247-3.131, p = .004.

## DISCUSSION

It is unclear why subjective memory concerns often persist long after concussion. The present study examined whether memory perfectionism, a core predisposing and/or perpetuating factor in FCD^19,20^, was associated with persistent memory complaints in a sample of adults seeking treatment post-concussion. Consistent with this hypothesis, we found that higher memory perfectionism was associated with severe memory complaints, after adjusting for objective memory, overall cognitive ability and depression. Sensitivity analyses suggest that this relationship is robust to the modality of objective memory measurement as well as to performance validity concerns. It does not appear that memory perfectionism is a function of higher post-concussion symptom reporting in general, as it did not predict severe fatigue complaint. In other words, memory perfectionism was specific in its relationship to elevated subjective memory concerns.

This is the first study to explore memory perfectionism in the context of post-concussion symptoms. Our findings are consistent with studies assessing people with FCD^21,22^. As in FCD, perceptions of poor memory following concussion may be fueled by excessive value and standards for one’s own memory abilities. Mechanistically, people with high memory perfectionism may have increased vigilance towards cognitive errors and anxiety about the permanence and consequences of brain injury. Heightened attention to minor memory lapses and distress-provoking interpretations of their meaning may further perpetuate memory complaints, and may represent a cognitive parallel of somatic hypervigilance^19,20^. People may also misremember having fewer memory lapses before their concussion than they actually did, an example of the “good-old-days” bias^34^. Additionally, post-injury stress, sleep problems, psychiatric disorders, and chronic pain may all contribute to both subjective and objective cognitive difficulties misattributed to brain injuy^8,9,12,35–38^.

There are several limitations to the current study. Memory perfectionism and memory complaints were assessed concurrently, at an average of four months post-injury. We therefore cannot be sure to what extent memory perfectionism is a premorbid personality trait and predisposing (i.e., vulnerability) factor or becomes (further) elevated after concussion. Relatedly, we cannot rule out reverse causality, that is, the possibility that people place importance on their memory ability and become intolerant of memory lapses *because* they experience memory problems. Our primary outcome was a single ordinal rating of how much participants’ experienced problems with forgetfulness now compared to before their injury. This item has demonstrated test-retest stability^39^ and strong discrimination between concussion and orthopedic injury control groups^40^, however, it may not have comparable reliability and validity to longer patient-reported outcomes assessing subjective memory functioning. Still, we believe that the presence vs. absence of severe memory complaints is a clinically relevant outcome. Participant characteristics that limit the generalizability of our findings include the high overall symptom burden (mean RPQ total score = 35.7) and advanced level of education (half had a post-secondary degree), which are not surprising for participants who are seeking treatment and willing to volunteer in research, but may not be representative of the broader concussion population. Our sample size limited us from considering additional covariates that may be important, such as anxiety and medications. Finally, it may be that performance on neuropsychological tests of memory are not sensitive enough to detect subtle real-world deficits that are reflected in people’s subjective experiences. Following this logic, relatively severe memory performance decrements (i.e., those detectable on neuropsychological testing) should be associated with more severe subjective memory problems. We and others^5–9^ have not observed this association.

In summary, the present study supports a role for memory perfectionism in persistent memory complaints after concussion. A full mechanistic model for persistent memory complaints awaits further validation and will likely include several other biopsychosocial pre- and post-injury factors. Subjective concern about memory decline, even in the absence of objective memory impairment, can result in functional impairment. For example, people may self-limit their participation in work or school after concussion due to low self-efficacy and anticipated negative consequences. Assessment of a patient with persistent memory complaints after concussion, especially when their symptoms are incompatible with their demonstrated memory ability on neuropsychological testing, should include exploration of their perceptions of their pre-injury memory ability, fears related to memory loss, reactions to memory lapses, and compensatory behaviors (e.g., reliance on others for help remembering things). Psychological interventions targeted at maladaptive beliefs about memory and counterproductive coping with perceived memory difficulties should be considered.

## Data Availability

All data produced in the present study are available upon reasonable request to the authors.

